# Cohort profile: The Pregnancy, Arsenic, and Immune Response (PAIR) Study, a longitudinal pregnancy and birth cohort in rural northern Bangladesh

**DOI:** 10.1101/2022.03.31.22273265

**Authors:** Lindsay N. Avolio, Tyler J. S. Smith, Ana Navas-Acien, Kate Kruczynski, Nora Pisanic, Pranay R. Randad, Barbara Detrick, Rebecca C. Fry, Alexander van Geen, Walter Goessler, Ruth A. Karron, Sabra L. Klein, Elizabeth L. Ogburn, Marsha Wills-Karp, Kelsey Alland, Kaniz Ayesha, Brian Dyer, Md. Tanvir Islam, Habibat A. Oguntade, Md. Hafizur Rahman, Hasmot Ali, Rezwanul Haque, Saijuddin Shaikh, Kerry J. Schulze, A. K. M. Muraduzzaman, A. S. M. Alamgir, Meerjady Sabrina Flora, Keith P. West, Alain B. Labrique, Christopher D. Heaney, the JiVitA Maternal and Child Health and Nutrition Research Project

## Abstract

**Purpose:** Arsenic exposure and micronutrient deficiencies may alter immune reactivity to influenza vaccination in pregnant women, transplacental transfer of maternal antibodies to the fetus, and maternal and infant acute morbidity. The Pregnancy, Arsenic, and Immune Response (PAIR) Study is a longitudinal pregnancy and birth cohort designed to assess whether arsenic exposure and micronutrient deficiencies alter maternal or newborn immunity and acute morbidity following maternal seasonal influenza vaccination during pregnancy.

**Participants:** We enrolled 784 pregnant women in rural Gaibandha District in northern Bangladesh between October 2018 and March 2019. Women received a quadrivalent seasonal inactivated influenza vaccine at enrollment in the late first or early second trimester between 11 and 17 weeks of gestational age. Follow-up included up to 13 visits between enrollment and three months postpartum as well as weekly telephone surveillance to ascertain influenza-like illness and other acute morbidity symptoms in women and infants. Tube well drinking water and urine specimens were collected to assess arsenic exposure. Of 784 women who enrolled, 736 (93.9%) delivered live births and 551 (70.3%) completed follow-up visit to three months postpartum.

**Findings to Date:** Arsenic was ≥0.02 µg/L in 97.9% of water specimens collected from participants at enrollment. The medians (interquartile ranges) of water and urinary arsenic were 5.1 (0.5-25.1) µg/L and 33.1 (19.6-56.5) µg/L, respectively. Water and urinary arsenic were strongly correlated (Spearman’s ρ=0.72) among women with water arsenic ≥ median but weakly correlated (ρ=0.18) among women with water arsenic < median.

**Future Plans:** The PAIR Study is well positioned to examine the effects of low-moderate arsenic exposure and micronutrient deficiencies on immune outcomes in women and infants.

**Registration:** NCT03930017

## INTRODUCTION

Arsenic exposure is a major threat to global health. About 140 million people worldwide are exposed to drinking water arsenic exceeding the World Health Organization’s (WHO’s) guideline value of 10 µg/L [1]. Arsenic causes bladder, lung, and skin cancers [2] and has been associated with cardiovascular disease, diabetes mellitus, and the metabolic syndrome [3,4]. Over the past decade, multiple studies have found that arsenic was associated with altered cellular [5] and humoral immune responses [6–8] and increased risk of infection, acute morbidity, and related mortality [9–12]. Of particular concern is immunotoxicity following exposure during pregnancy and early life [13]. However, while exposure *in utero* has been associated with reduced pathogen-specific antibody responses to some childhood vaccinations [6–8] and increased risk of respiratory and gastrointestinal morbidities in children [14–21], less is known about arsenic and the immune response in pregnant women and newborns during the first months of life. In addition, arsenic methylation facilitated by one-carbon metabolism appears to modify arsenic toxicity for certain chronic disease outcomes [22]. Arsenic methylation is known to shift during pregnancy [23,24]. However, few studies of arsenic immunotoxicity have evaluated potential effect measure modification by micronutrient deficiencies that influence arsenic methylation in pregnant women.

The WHO recommends seasonal influenza vaccination at any stage of pregnancy to protect pregnant women and infants <6 months of age [25], who benefit from maternal antibodies transferred across the placenta [26–28]. Since the risk of severe illness from infection by influenza virus is higher in pregnant women and infants [29,30], expanding maternal vaccination against influenza is imperative. If arsenic reduces the maternal antibody response to influenza vaccine or transplacental transfer of maternal antibodies to the fetus, however, additional interventions may be needed. Yet relations among arsenic exposure, micronutrient deficiencies, antibody responses to influenza vaccination in pregnant women, transplacental transfer of maternal antibodies, and influenza-like illness (ILI) and other acute morbidities remain understudied [31,32].

To better understand the influence of arsenic on immune responses in pregnant women and newborns, we established the Pregnancy, Arsenic, and Immune Response (PAIR) Study, a longitudinal pregnancy and birth cohort, in rural northern Bangladesh. The PAIR Study was designed to assess whether arsenic exposure and micronutrient deficiencies alter maternal or newborn immunity and acute morbidity following maternal seasonal influenza vaccination during pregnancy. We hypothesized that arsenic exposure and one-carbon metabolism micronutrient deficiencies may alter maternal and newborn influenza antibody titer and avidity, measures of systemic immune responses, and respiratory and other acute morbidity outcomes among pregnant women and newborns.

## COHORT DESCRIPTION

### Study Area

The PAIR Study is based at the JiVitA Maternal and Child Health and Nutrition Research Project (JiVitA) in rural northern Bangladesh. Approximately 45 million people in Bangladesh are exposed to arsenic concentrations in drinking water >10 µg/L (the WHO guideline value) and, of these, approximately 20 million people are exposed to arsenic concentrations >50 µg/L (the Bangladesh national standard) [33]. For nearly two decades, JiVitA has been one of the largest research project sites in South Asia, covering ∼650,000 people over 450 km^2^ of Bangladesh’s Gaibandha and Rangpur Districts (**Figure 1**) [34]. JiVitA is an extensive field organization that has conducted large trials of food and micronutrient supplements among pregnant women and children [35–37]. A case-control study nested in a previous JiVitA micronutrient supplement trial found that urinary arsenic was associated with seroconversion to hepatitis E virus between the first trimester and three months postpartum among pregnant women in the study area [9]. The PAIR Study builds on this work.

**Figure 1.**
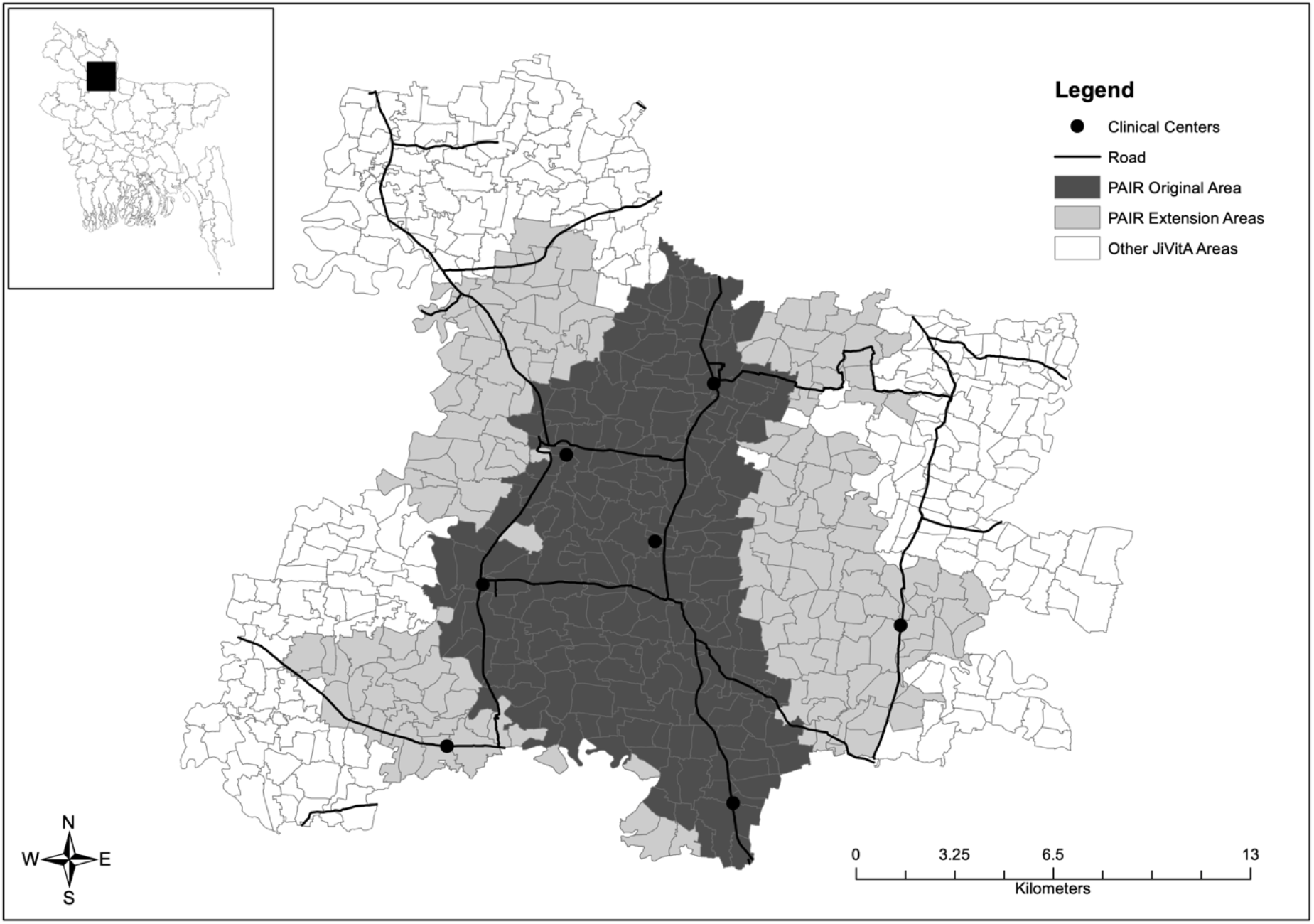
Map of the Pregnancy, Arsenic, and Immune Response (PAIR) Study area within the JiVitA Maternal and Child Health and Nutrition Research Project in northern Bangladesh, indicating the original study area (dark gray), extensions to the study area to achieve the target sample size in the same influenza season and before the Ramadan fast (light gray), and remaining JiVitA area (white). Sector boundaries are represented by light gray lines. Clinical centers where biospecimen collection was performed are noted by black circles. Major roads are indicated by black lines. The inset indicates the approximate location of the PAIR Study (black square) in Bangladesh.

### Enrollment and Vaccination

From July 2018 to February 2019, we conducted pregnancy surveillance to identify eligible pregnant women in the study area. A married woman of reproductive age (13-45 years) was eligible for the PAIR Study if she was pregnant and 13-14 weeks of gestational age (GW) (later 13-16 GW; see below), had no pre-existing immunodeficiency or chronic infection, had no previous or current use of immune-altering drugs or therapies (*e*.*g*., steroids), and had not yet received an influenza vaccine for the 2018-19 influenza season.

The full JiVitA study area is divided into 566 sectors of ∼150-300 households each (**Figure 1**). For the PAIR Study, surveillance began in 175 sectors closest to clinical centers where vaccination and biospecimen collections, including venipuncture, could be safely performed and samples could be uniformly processed (“PAIR Original Area” in **Figure 1**). We estimated *a priori* that enrolling 850 pregnant women in 13-14 GW would yield a target sample size of 400 mother-infant pairs retained in the study to three months postpartum. We also determined *a priori* that enrollment should end by March 2019 so that pregnancies would be limited to a single influenza season and follow-up visits at 28 days post-vaccination would occur before the Ramadan fast (May 5 to June 3, 2019). In November 2018, seeking to achieve the target enrollment in this time window, we enlarged the study area by 163 sectors (“PAIR Extension Areas” in **Figure 1**) and expanded eligibility criteria to 13-16 GW.

A total of 66,055 married women of reproductive age living in the original or extension areas were identified by an earlier census or during pregnancy surveillance (**Figure 2**). During surveillance, a female community health research worker (CHRW) visited each household every four weeks and asked each married woman of reproductive age about the date of her last menstrual period. If the date was >30 days prior to the visit, the woman was offered a urine pregnancy test. A positive test indicated that the woman was pregnant. Gestational age was calculated from the date of her last menstrual period. We completed at least one surveillance visit to 46,775 women to identify 2,615 pregnant women (**Figure 2**). Of these, 845 women were eligible and consented to enrollment, and 784 women enrolled in the PAIR Study between October 2018 and March 2019 (**Figure 2**). At enrollment, women received a quadrivalent seasonal inactivated influenza vaccine (VaxigripTetra, Sanofi Pasteur, Lyon, France), which was recommended by the WHO for the 2018-19 Northern Hemisphere influenza season. We enrolled and administered the vaccine to 8 women in GW 11-12, 775 women in GW 13-16, and 1 woman in GW 17. All vaccines were maintained under a cold-chain protocol, carefully temperature-monitored, and administered by trained nurses working under the supervision of a JiVitA physician. The vaccine contained four inactivated influenza virus strains: A/Michigan/2015 (H1N1), A/Singapore/2016 (H3N2), B/Maryland/2016, and B/Phuket/2013 [38].

**Figure 2.**
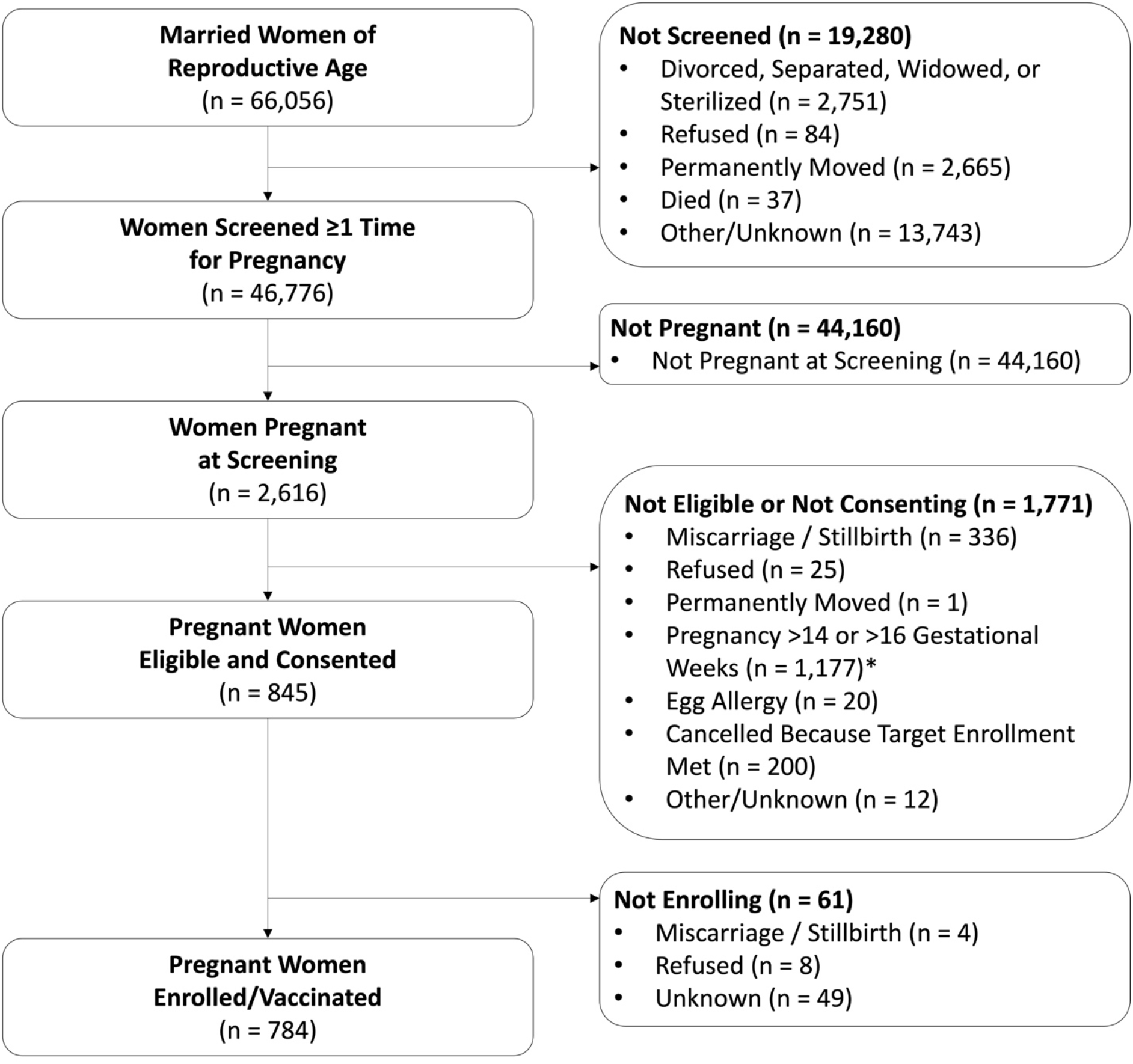
Eligibility and enrollment in the Pregnancy, Arsenic, and Immune Response (PAIR) Study, Gaibandha District, Bangladesh, 2018-2019. We conducted pregnancy surveillance from July 2018 to February 2019 and enrolled eligible pregnant women from October 2018 to March 2019. *Initially, pregnant women were eligible at 13-14 weeks of gestational age (GW). In November 2018, we expanded eligibility to 13-16 GW.

Sociodemographic characteristics, including age, education, socioeconomic status, household size (number of people), and house size (number of rooms excluding the kitchen and storerooms), were similar between pregnant women who enrolled (n=784) and pregnant women who did not enroll in the PAIR Study (n=1,519, after excluding women who were missing sociodemographic information [n=313]) (**Table 1**). Women who enrolled tended to be younger and have higher socioeconomic status, but the differences were small (**Table 1**). Socioeconomic status was assessed by living standards index, which was generated by a principal components analysis of household assets and home construction materials and validated in a previous JiVitA study [39]. The use of tube wells to obtain groundwater for drinking and cooking was nearly universal among enrollees (99.4%) and non-enrollees (99.8%) alike (**Table 1**).

**Table 1.** Sociodemographic characteristics [n (%)] of pregnant women in the study area by enrollment in the Pregnancy, Arsenic, and Immune Response (PAIR) Study, Gaibandha District, Bangladesh, 2018-2019.

|  | **Enrolled**<br>(n=784) | **Not Enrolled\***<br>(n=1,519) | *p* |
| --- | --- | --- | --- |
| **Age (years)\*\*** |  |  |  |
| <20 | 89 (11.4) | 175 (11.5) | <0.01 |
| 20 to 29 | 499 (63.6) | 859 (56.6) |  |
| ≥30 | 194 (24.7) | 479 (31.5) |  |
| Missing | 2 (0.3) | 6 (0.4) |  |
| **Maternal Education** |  |  |  |
| None | 92 (11.7) | 212 (14.0) | 0.14 |
| Class 1 to 9 | 548 (69.9) | 1,065 (70.1) |  |
| Class ≥10 | 144 (18.4) | 240 (15.8) |  |
| Missing | 0 (0) | 2 (0.1) |  |
| **Living Standards Index** |  |  |  |
| <Median | 360 (45.9) | 792 (52.1) | <0.01 |
| ≥Median | 424 (54.1) | 727 (47.9) |  |
| **Household Size (people)** |  |  |  |
| 2 to 3 | 307 (39.2) | 518 (34.1) | 0.05 |
| 4 to 5 | 336 (42.9) | 720 (47.4%) |  |
| ≥6 | 139 (17.7) | 277 (18.2) |  |
| Missing | 2 (0.3) | 4 (0.3) |  |
| **House Size (rooms)\*\*\*** |  |  |  |
| 0 to 1 | 366 (46.7) | 721 (47.5) | 0.93 |
| 2 to 3 | 367 (46.8) | 699 (46.0) |  |
| ≥4 | 51 (6.5) | 98 (6.5) |  |
| Missing | 0 (0) | 1 (0.1) |  |
| **Drinking Water Source** |  |  |  |
| Tube Well | 779 (99.4) | 1,516 (99.8) | - |
| Other | 5 (0.6) | 3 (0.2) |  |

*Notes: All p values were calculated using $\chi^2$ tests. A p value for Drinking Water Source was omitted because the number of people in the "Other" category was too small. \*The table does not include non-enrolled women who did not complete a separate visit to assess sociodemographic variables (n=313). \*\*Age as of July 2018 (start of pregnancy surveillance). \*\*\*Number of rooms excluding kitchen and storerooms.*

### Follow-up

#### Study Visits

##### In-person Visits

We completed up to 13 in-person visits to implement questionnaires and collect biological and environmental specimens, beginning at enrollment and continuing to three months postpartum, for a follow-up period of roughly 10-11 months, depending on the timing and duration of each pregnancy (**Tables 2 and 3**). These included up to eight visits to the woman during pregnancy and up to five visits to mother-infant pairs after live birth. At three visits (enrollment and vaccination, 28 days post-vaccination, and three months postpartum), the CHRW brought participants (the mother in pregnancy, the mother-infant pair after live birth) to a local clinical center for detailed questionnaire and biospecimen collection. To minimize burden on mothers and infants in the neonatal period, two home visits were conducted shortly after any live birth and again within the first month postpartum for further questionnaire and biospecimen collection. Of 784 women who enrolled, 744 (94.9%) completed the 28-day post-vaccination visit, 598 (76.3%) completed a visit within the first month postpartum, and 551 (70.3%) completed the three-month postpartum visit (**Table 4**). In total, 736 (93.9%) enrolled women had 750 live births. Age, gestational age, parity, education, living standards index, household size (number of people), house size (number of rooms), height (cm), body mass index (kg/m^2^), and urinary arsenic were similar among the women who enrolled and the subset of women who completed follow-up at three months postpartum (**Table 4**).

**Table 2.**
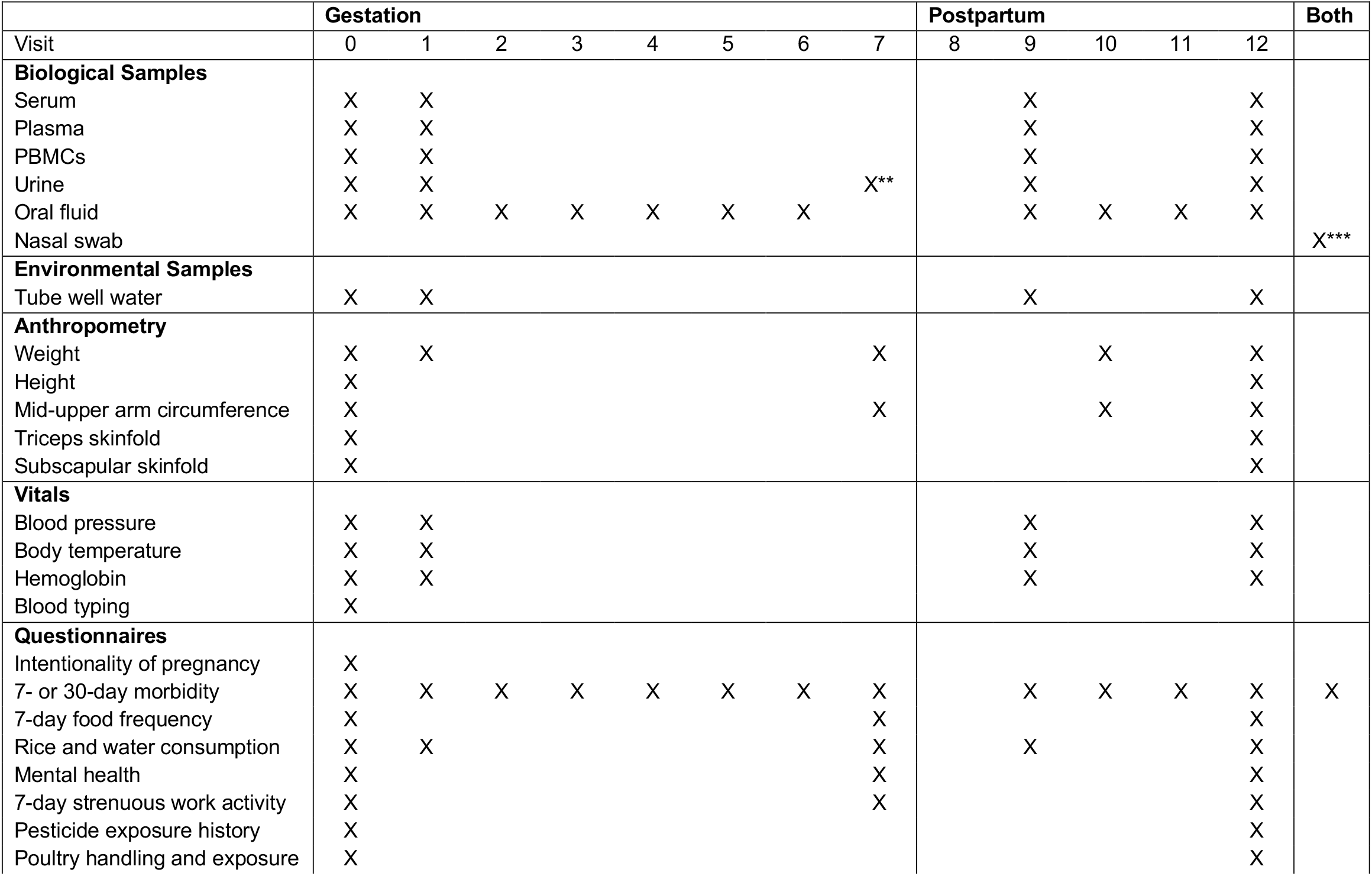
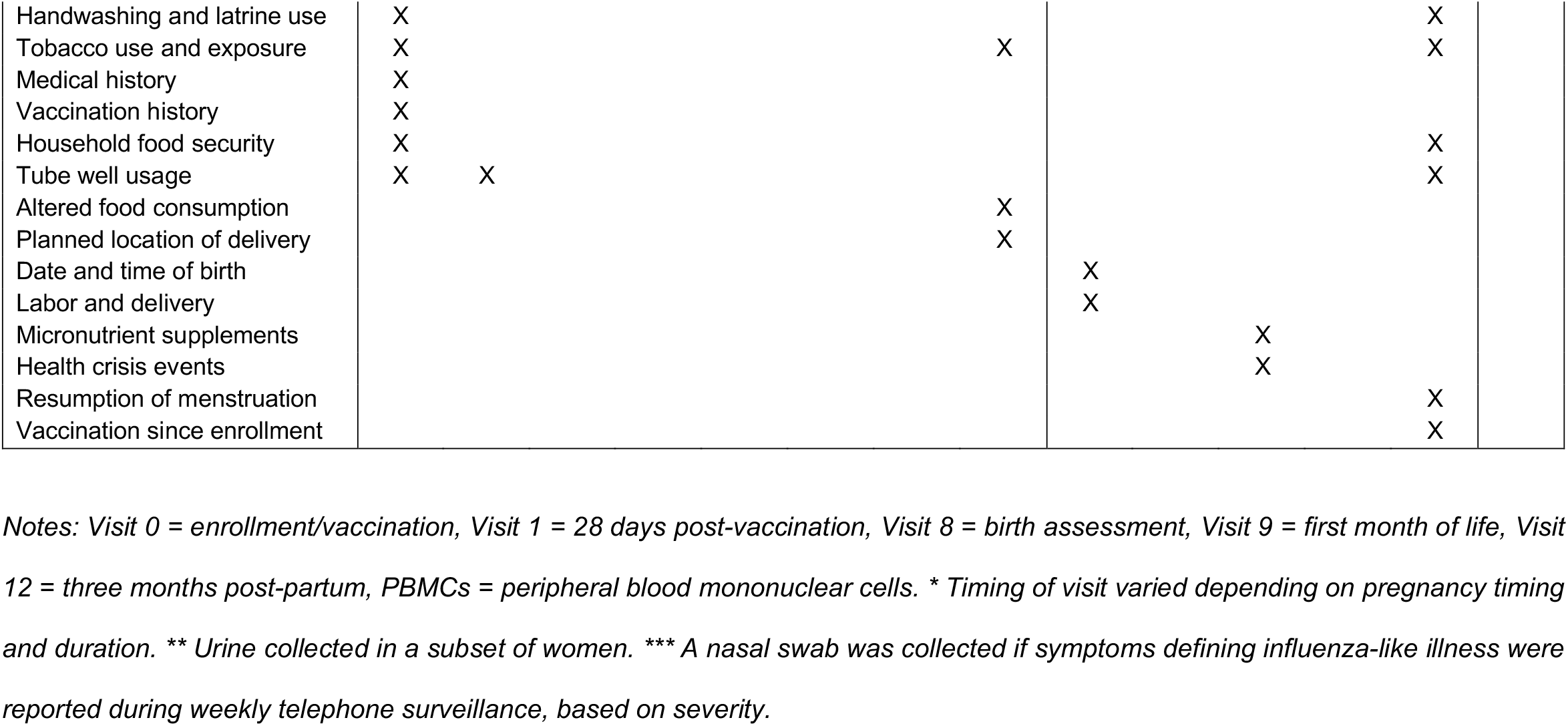
Measurements collected in pregnant women in the Pregnancy, Arsenic, and Immune Response (PAIR) Study, Gaibandha District, Bangladesh, 2018-2019.

**Table 3.** Measurements collected in infants in the Pregnancy, Arsenic, and Immune Response (PAIR) Study, Gaibandha District, Bangladesh, 2018-2019.

|  | Postpartum |  |  |  |  | Both |
| --- | --- | --- | --- | --- | --- | --- |
| Visit | 8 | 9 | 10 | 11 | 12 |  |
| **Biological Samples** |  |  |  |  |  |  |
| Serum |  | X |  |  | X |  |
| Oral fluid |  |  | X | X | X |  |
| Nasal swab |  |  |  |  |  | X* |
| **Anthropometry** |  |  |  |  |  |  |
| Weight | X |  | X |  | X |  |
| Length | X |  | X |  | X |  |
| Mid-upper arm circumference | X |  | X |  | X |  |
| Chest circumference | X |  | X |  | X |  |
| Head circumference | X |  | X |  | X |  |
| **Vitals** |  |  |  |  |  |  |
| Body temperature |  | X |  |  | X |  |
| Hemoglobin |  |  |  |  | X |  |
| Blood typing |  |  |  |  | X |  |
| **Questionnaires** |  |  |  |  |  |  |
| Date and time of birth | X |  |  |  |  |  |
| Birth outcome | X |  |  |  |  |  |
| Condition of infant at birth | X |  |  |  |  |  |
| Newborn care practices | X |  |  |  |  |  |
| 7-day morbidity |  | X | X | X | X | X |
| Morbidities in first 28 days |  |  | X |  |  |  |
| Breastfeeding practices |  |  | X |  | X |  |
| Early feeding |  |  | X |  |  |  |
| Injury and bleeding |  |  | X |  |  |  |
| Two-month morbidity | X |  |  |  |  |  |
| Child's diet | X |  |  |  |  |  |
| Immunization history |  |  |  |  | X |  |
| Vitamin A supplementation |  |  |  |  | X |  |
| Bleeding disorders |  |  |  |  | X |  |
| Hospitalization history |  |  |  |  | X |  |
| Recent medication |  |  |  |  | X |  |

*Notes: Visit 8 = birth assessment, Visit 9 = first month of life, Visit 12 = three months post-partum.*
*\* A nasal swab was collected if symptoms defining influenza-like illness were reported during weekly telephone surveillance, based on severity.*

**Table 4.**
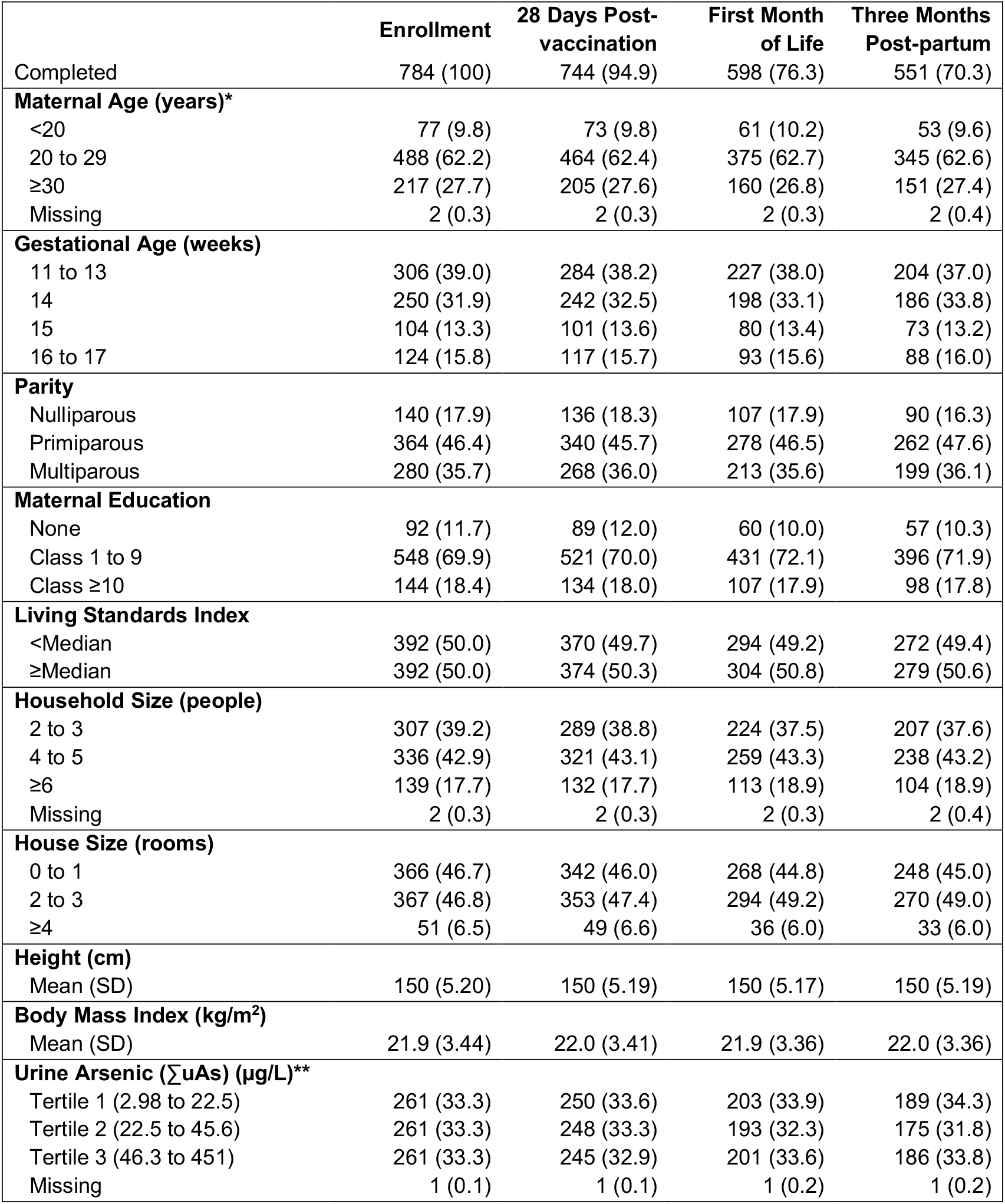
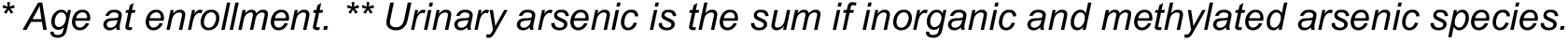
Sociodemographic characteristics [n (%)] of pregnant women at enrollment in the Pregnancy, Arsenic, and Immune Response (PAIR) Study, Gaibandha District, Bangladesh, 2018-2019, by completion of major study visits.

|  | **Enrollment** | **28 Days Post-vaccination** | **First Month of Life** | **Three Months Post-partum** |
| --- | --- | --- | --- | --- |
| Completed | 784 (100) | 744 (94.9) | 598 (76.3) | 551 (70.3) |
| **Maternal Age (years)\*** |  |  |  |  |
| <20 | 77 (9.8) | 73 (9.8) | 61 (10.2) | 53 (9.6) |
| 20 to 29 | 488 (62.2) | 464 (62.4) | 375 (62.7) | 345 (62.6) |
| ≥30 | 217 (27.7) | 205 (27.6) | 160 (26.8) | 151 (27.4) |
| Missing | 2 (0.3) | 2 (0.3) | 2 (0.3) | 2 (0.4) |
| **Gestational Age (weeks)** |  |  |  |  |
| 11 to 13 | 306 (39.0) | 284 (38.2) | 227 (38.0) | 204 (37.0) |
| 14 | 250 (31.9) | 242 (32.5) | 198 (33.1) | 186 (33.8) |
| 15 | 104 (13.3) | 101 (13.6) | 80 (13.4) | 73 (13.2) |
| 16 to 17 | 124 (15.8) | 117 (15.7) | 93 (15.6) | 88 (16.0) |
| **Parity** |  |  |  |  |
| Nulliparous | 140 (17.9) | 136 (18.3) | 107 (17.9) | 90 (16.3) |
| Primiparous | 364 (46.4) | 340 (45.7) | 278 (46.5) | 262 (47.6) |
| Multiparous | 280 (35.7) | 268 (36.0) | 213 (35.6) | 199 (36.1) |
| **Maternal Education** |  |  |  |  |
| None | 92 (11.7) | 89 (12.0) | 60 (10.0) | 57 (10.3) |
| Class 1 to 9 | 548 (69.9) | 521 (70.0) | 431 (72.1) | 396 (71.9) |
| Class ≥10 | 144 (18.4) | 134 (18.0) | 107 (17.9) | 98 (17.8) |
| **Living Standards Index** |  |  |  |  |
| <Median | 392 (50.0) | 370 (49.7) | 294 (49.2) | 272 (49.4) |
| ≥Median | 392 (50.0) | 374 (50.3) | 304 (50.8) | 279 (50.6) |
| **Household Size (people)** |  |  |  |  |
| 2 to 3 | 307 (39.2) | 289 (38.8) | 224 (37.5) | 207 (37.6) |
| 4 to 5 | 336 (42.9) | 321 (43.1) | 259 (43.3) | 238 (43.2) |
| ≥6 | 139 (17.7) | 132 (17.7) | 113 (18.9) | 104 (18.9) |
| Missing | 2 (0.3) | 2 (0.3) | 2 (0.3) | 2 (0.4) |
| **House Size (rooms)** |  |  |  |  |
| 0 to 1 | 366 (46.7) | 342 (46.0) | 268 (44.8) | 248 (45.0) |
| 2 to 3 | 367 (46.8) | 353 (47.4) | 294 (49.2) | 270 (49.0) |
| ≥4 | 51 (6.5) | 49 (6.6) | 36 (6.0) | 33 (6.0) |
| **Height (cm)** |  |  |  |  |
| Mean (SD) | 150 (5.20) | 150 (5.19) | 150 (5.17) | 150 (5.19) |
| **Body Mass Index (kg/m<sup>2</sup>)** |  |  |  |  |
| Mean (SD) | 21.9 (3.44) | 22.0 (3.41) | 21.9 (3.36) | 22.0 (3.36) |
| **Urine Arsenic (ΣuAs) (μg/L)\*\*** |  |  |  |  |
| Tertile 1 (2.98 to 22.5) | 261 (33.3) | 250 (33.6) | 203 (33.9) | 189 (34.3) |
| Tertile 2 (22.5 to 45.6) | 261 (33.3) | 248 (33.3) | 193 (32.3) | 175 (31.8) |
| Tertile 3 (46.3 to 451) | 261 (33.3) | 245 (32.9) | 201 (33.6) | 186 (33.8) |
| Missing | 1 (0.1) | 1 (0.1) | 1 (0.2) | 1 (0.2) |

\* Age at enrollment. \*\* Urinary arsenic is the sum of inorganic and methylated arsenic species.

##### Weekly Telephone Surveillance

In partnership with the Institute of Epidemiology Disease Control and Research (IEDCR), a unit of the Bangladesh Ministry of Health and Family Welfare, we conducted weekly telephone surveillance to ascertain acute morbidities in women (enrollment to three months postpartum) and infants (birth to three months of age). In women, we ascertained high fever with cough, high fever with sore throat, diarrhea, vomiting, and abdominal pain. If a woman reported high fever with cough or sore throat, she was asked further if she had congestion, headache, or chills at the same time. In infants, we ascertained high fever with cough, diarrhea, and vomiting. If a woman reported that her infant had high fever with cough, she was asked further if the infant had congestion or shortness of breath at the same time. Of 784 women enrolled in the PAIR Study and 750 infants born to them, 743 women (94.8%) and 628 infants (83.7%) participated in the surveillance. While weekly telephone surveillance was prespecified to end at three months postpartum, for some women and infants it continued for longer. The median (interquartile range [IQR]) number of weeks with complete calls was 35 (23-42) for women and 17 (12-22) for infants.

#### Environmental Specimens

We collected a water specimen from the tube well that each participant indicated was her primary source of drinking water before each center visit and at the home visit conducted within one month postpartum (**Table 2**). Just prior to collection, the well was flushed for approximately five minutes. Specimens were collected into conical vials certified by the manufacturer as trace-metal free (<1 µg/L for 20 metals, including As), and transported to the local JiVitA lab at 4-10°C. Specimens were aliquoted and stored at -20°C prior to analysis. Tube well water specimens were analyzed for total arsenic (wAs) and other elements (Al, Ba, Br, Ca, Cd, Cu, Fe, K, Mg, Mn, Mo, Na, P, Pb, S, Sb, Si, Sr, U, V, W, Zn) at the Lamont-Doherty Earth Observatory in Palisades, NY using inductively coupled plasma mass spectrometry [40]. Empty trace metal-free conical vials and aliquot tubes were assessed as blanks. The LOD for wAs was 0.02 µg/L.

#### Biological Specimens

Up to 22 mL of venous blood was collected from mothers at each of the three center visits: enrollment/vaccination, 28 days post-vaccination, and three months postpartum (**Table 2**). Up to 1 mL of capillary blood was collected from infants by heel stick during the center visit at three months postpartum (**Table 3**). At the home visit within approximately one month of live birth, blood was collected from mothers and infants using the methods described for the center visits. Blood specimens were transported in temperature-controlled and -monitored coolers to a JiVitA laboratory. Laboratory technicians processed maternal blood to serum, plasma, and peripheral blood mononuclear cells (PBMCs), and processed infant blood to serum, on the day of collection. Serum and plasma were frozen and maintained at -80°C. PBMCs were cryopreserved following established protocols [41]. Serum and plasma were shipped on dry ice, and PBMCs were shipped in vapor phase liquid nitrogen canisters, to the Johns Hopkins University in Baltimore, Maryland. Maternal and infant sera were tested for antibody for the influenza vaccine antigens by hemagglutination-inhibition (HAI) assay at Sanofi Pasteur in Swiftwater, Pennsylvania [42]. We also will determine avidity of IgG antibodies specific to the influenza vaccine antigens; serum cytokine and chemokine concentrations as measures of immune function; immune cell population characterization; and T-cell stimulation assays using the influenza vaccine antigens. Micronutrients relevant to one-carbon metabolism will be measured in maternal plasma.

Urine was collected from all mothers at the three center visits and during the home visit within one month of birth, and from a subset of mothers (n=468) during a home visit in late pregnancy (**Table 2**). Mothers were asked to provide a urine specimen in a collection cup. A CHRW immediately transferred the specimen to a container certified by the manufacturer as trace-metal free (<1 µg/L for 20 metals, including As) and the specimens were transported to the JiVitA laboratory. Urine specimens were aliquoted and stored at -20°C prior to analysis. Urinary elements (Al, Ba, Br, Ca, Cd, Cs, Cu, Fe, K, Li, Mg, Mn, Mo, Na, P, Pb, Rb, S, Sb, Se, Si, Sr, U, V, W, Zn) were measured at the Institute of Chemistry - Analytical Chemistry at the University of Graz in Graz, Austria using inductively coupled plasma tandem mass spectrometry (ICP-MS/MS). Urinary arsenic was speciated (arsenite, arsenate, monomethylarsonic acid (MMA), dimethylarsinic acid (DMA), and arsenobetaine and other cations) by high performance liquid chromatography (HPLC) with ICP-MS/MS as described by Scheer et al. [43]. Empty trace-metal free containers and aliquot tubes were assessed as blanks. The limits of detection (LODs) for arsenic species were 0.05 µg/L. To assess arsenic exposure, we summed urinary inorganic arsenic (the sum of arsenite and arsenate; iAs), MMA, and DMA (∑uAs) [44]. To assess arsenic metabolism, we calculated proportions of urinary iAs, MMA, and DMA, relative to ∑uAs, and reported them as iAs%, MMA%, and DMA% [22]. All urinary concentrations were corrected for specific gravity measured by refractometric determination of total solids.

We collected oral fluid, which includes saliva, oral mucosal transudate from the capillary bed, and crevicular fluid, from mothers at each center visit, the home visit after birth, and additional monthly home visits, and from infants in all visits beginning at one month postpartum (**Tables 2 and 3**). Crevicular fluid, which flows between the gums and the teeth, is rich in antibodies and reflects the IgG profile of the serum. Oral fluid was collected by brushing the gum line with an oral-mucosal transudate collection swab [45–47]. Microsphere magnetic bead-based assays will be applied to measure IgG and IgA responses specific to the influenza vaccine strains as well as a variety of endemic respiratory and gastrointestinal pathogens [45,46,48].

If a mother reported influenza-like illness (ILI) in herself or her infant during weekly surveillance, a CHRW aimed to conduct a home visit and collect a nasal swab within 24 hours of the call (**Tables 2 and 3**). ILI was defined as high fever with cough or sore throat in women and high fever with cough in infants [49]. Severity was determined by calculating the sum of acute morbidity symptoms reported during surveillance. Given limited capacity, women or infants with the highest sum score were prioritized for the collection of a nasal swab. After collection by a CHRW, swabs were placed in universal transport medium. The transport medium was intended for the preservation of viruses (Puritan UniTranz-RT, Guilford, ME, USA). Samples were transported to the local JiVitA lab and stored at -80°C. Nasal swabs were periodically batch-transported to IEDCR in Dhaka, Bangladesh, in liquid nitrogen, where RNA will be extracted and tested by RT-qPCR for influenza A and B virus, including sub-types, according to WHO protocols to assess laboratory-confirmed ILI [50].

#### Anthropometry

Maternal weight (kg), height (cm), mid-upper arm circumference (MUAC) (cm), and triceps and subscapular skinfolds (mm) were measured at enrollment/vaccination and three months postpartum (**Table 2**) [36,43,52]. Maternal weight was also measured at 28 days post-vaccination, late pregnancy, and one-month postpartum. Maternal MUAC was also measured at late pregnancy and one-month postpartum. Infant weight, MUAC, chest circumference, head circumference, and length were recorded at the birth assessment visit (**Table 3**) [36,43,52]. Infant growth was monitored by repeating these measurements at one- and three-months postpartum.

#### Vitals

Blood pressure, hemoglobin, and body temperature of mothers were measured by a local nurse each time a mother had blood drawn (**Table 2**). Systolic and diastolic blood pressure were measured twice, in the left arm, with the woman in a relaxed and seated position using a WelchAllyn DuraShock DS66 Trigger Aneroid Sphygmomanometer (Skaneateles Falls, New York, USA). Hemoglobin was measured in capillary blood by real-time assay using a HemoCue Hb 301 analyzer (Ängelholm, Sweden). Body temperature of infants was measured at the birth assessment and three months postpartum, and hemoglobin was measured at three months postpartum by a local nurse (**Table 3**). Vitals were recorded and provided to the mother after each collection. Pregnant women and infants who were severely anemic (hemoglobin <7 g/dL)[53] were given iron tablets and iron syrup, respectively, consistent with the study protocol and as approved by the IRB. Blood type was determined using anti-A, anti-B, and anti-D blood grouping reagents (Cromatest, LiNEAR, Montgat, Spain).

#### Questionnaires

Questionnaires conducted at multiple visits between enrollment/vaccination and three months postpartum covered a variety of topics (**Table 2**), including questions related to the intentionality of pregnancy, seven- and thirty-day morbidity including influenza-like illness, seven-day food frequency, mental health, seven-day strenuous work activity, six-month pesticide storage and use, poultry farming and exposures, handwashing practices, tobacco use and smoke exposure, medical and vaccination history, household food security, and more. Additional questionnaires asked about the birth and delivery of infants, including characteristics of labor and delivery, micronutrient supplement consumption, and health crisis events. Answers to questionnaires for infants were most often provided by the mother, and included morbidity, breastfeeding practices, early feeding practices, infant injury and bleeding, immunization and supplementation, and hospitalization history and medication use (**Table 3**). Topics related to arsenic exposure included drinking water, rice products, and tobacco products and smoke exposure in the previous week.

### Patient and Public Involvement

JiVitA was established and operates in consultation with local community stakeholders, many of whom are hired as JiVitA staff. JiVitA has benefited from their continued support for nearly two decades. Participants were not directly involved in the design of the PAIR Study.

### FINDINGS TO DATE

Water arsenic was ≥0.02 µg/L in 97.9% of drinking water specimens collected from participants’ tube wells on the day of enrollment. The median (IQR) of wAs was 5.1 (0.5-25.1) µg/L (**Figure 3**) with 39.4% of samples above the WHO guideline value of 10 µg/L and 13.2% above the Bangladesh national standard of 50 µg/L. wAs appears to be lower in the study area than in other regions of Bangladesh where epidemiologic studies have been conducted [54,55]. Urinary iAs, MMA, and DMA were ≥LOD in all specimens collected at enrollment. The median (IQR) of their sum, ∑uAs, was 33.1 (19.6-56.5) µg/L (**Figure 3**). Urinary arsenobetaine (and other unretained arsenic species) levels were low (median: 0.8 µg/L, IQR: 0.4-1.7 µg/L), suggesting low exposure to organic arsenic from seafood in the population [56]. ∑uAs was strongly correlated with wAs in participants with wAs ≥ median (Spearman’s ρ=0.72) but weakly correlated with wAs in participants with wAs < median (Spearman’s ρ=0.18) (**Figure 4**). This suggests that, at lower concentrations of wAs, other sources of arsenic (*e*.*g*., other tube wells, rice) contribute relatively more to exposure [57]. ∑uAs and wAs will provide complementary approaches to exposure assessment. ∑uAs integrates exposures from multiple sources, including drinking water and diet, but has a short biological half-life and may over- or under-estimate long-term exposure [44]. By contrast, wAs reflects a single source of exposure but is more stable over time. Future work will characterize exposure to arsenic and other metals and associations with health outcomes, including maternal antibody response to influenza vaccination, transplacental transfer of maternal antibodies, and respiratory morbidity, as well as interactions between arsenic exposure and micronutrient deficiencies in these outcomes.

**Figure 3.**
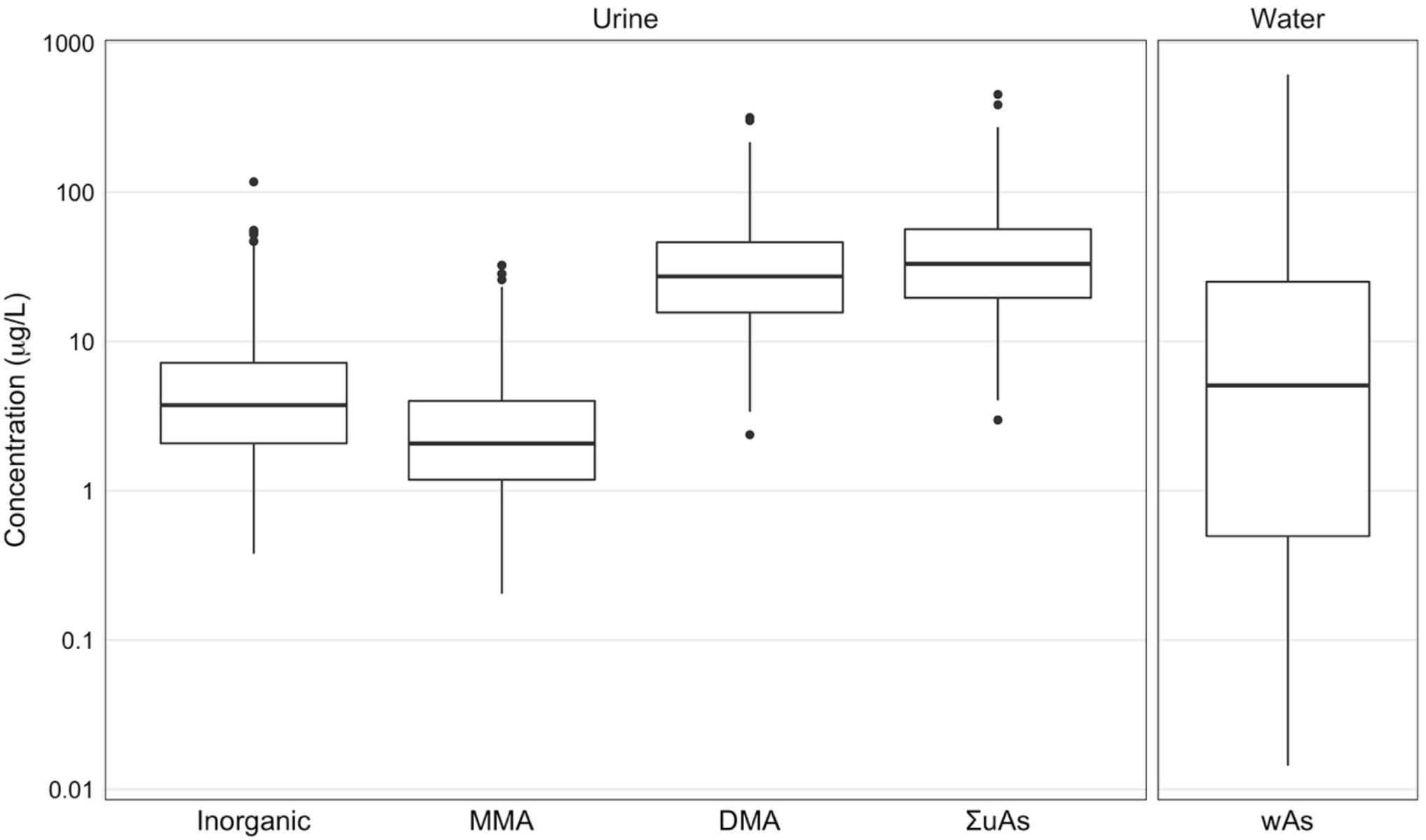
Tube well water arsenic (wAs) and urinary inorganic, monomethyl (MMA), and dimethyl (DMA) arsenic, and their sum (∑uAs), plotted on a common log scale, among pregnant women at enrollment in the Pregnancy, Arsenic, and Immune Response (PAIR) Study, Gaibandha District, Bangladesh, 2018-2019.

**Figure 4.**
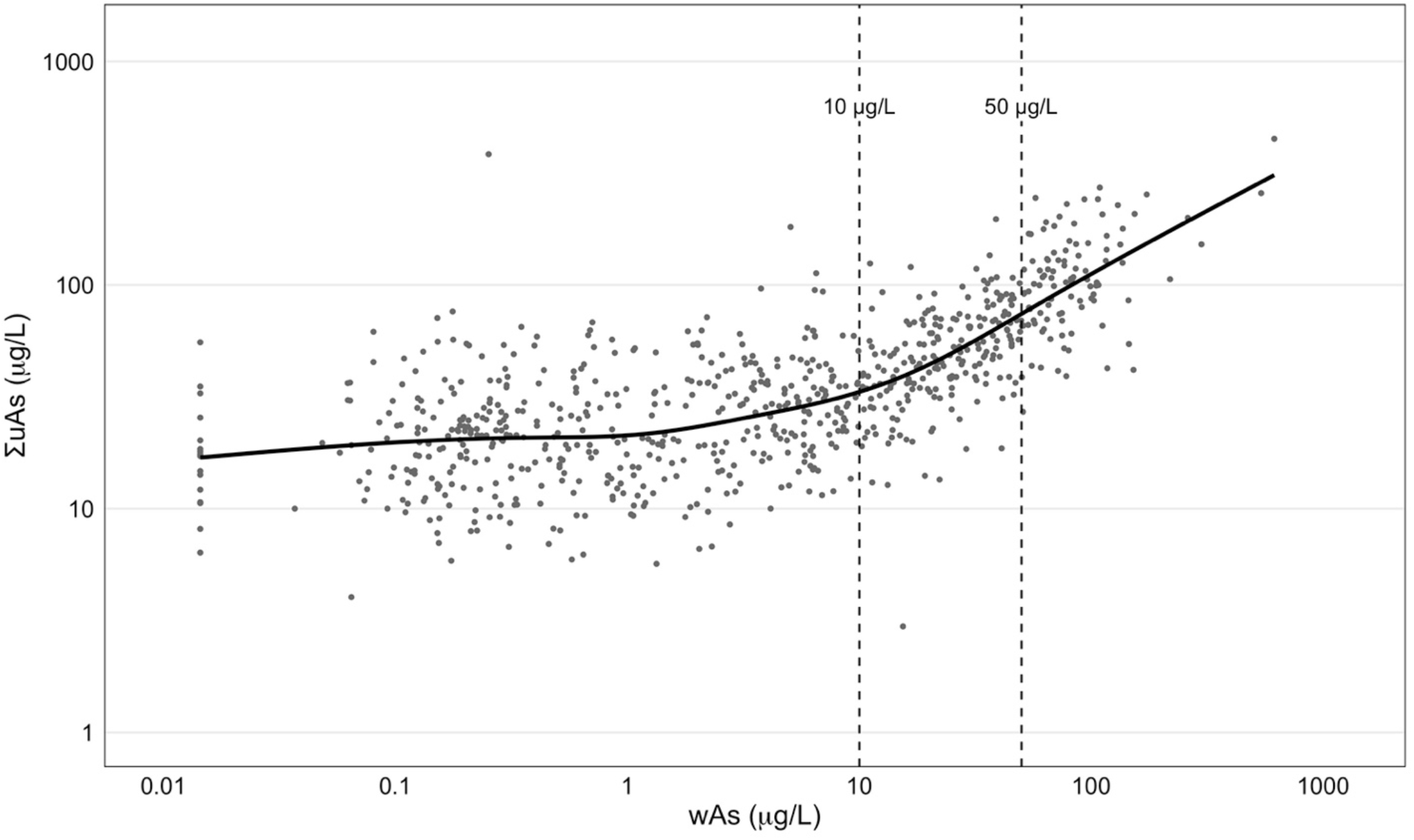
Tube well water arsenic (wAs) and the sum of urinary inorganic, monomethyl, and dimethyl arsenic (∑uAs), plotted on a common log scale, among pregnant women at enrollment in the Pregnancy, Arsenic, and Immune Response (PAIR) Study, Gaibandha District, Bangladesh, 2018-2019. A generalized additive model smoother is shown by the solid line. Drinking water arsenic standards for the World Health Organization (10 µg/L) and Bangladesh (50 µg/L) are indicated by dashed vertical lines.

### STRENGTHS AND LIMITATIONS

Strengths of the PAIR Study include a large, representative sample of pregnant women and infants in rural northern Bangladesh. The women received the same seasonal influenza vaccine at approximately the same time during pregnancy and during the same influenza season. This design will enable us to assess whether arsenic exposure is associated with antibody responses to seasonal influenza vaccination in pregnant women or transplacental transfer of maternal antibodies to the fetus while avoiding potential confounders related to stage of pregnancy and annual variability in influenza viruses. Because wAs appears to be lower in the PAIR Study than in other epidemiologic studies in Bangladesh, we can assess whether low-moderate arsenic exposure is associated with relevant maternal and infant outcomes. Biological and environmental specimens provide objective measures of arsenic exposure and immune and respiratory and other acute morbidity outcomes. Weekly telephone surveillance provides ascertainment of ILI symptoms and real-time PCR testing of ILI-symptomatic event anterior nasal swabs provides time-resolved information on maternal and infant laboratory-confirmed influenza A/B infection. Limitations include occasional delays in study visits. While JiVitA possesses an extensive field organization, logistical barriers to conducting frequent home visits over a large study area prevented some study visits from occurring as scheduled. Additionally, it was not logistically feasible to collect umbilical cord blood specimens to assess transplacental transfer of maternal antibodies. However, infant-to-maternal HAI antibody titer ratios measured in serum during the first month of life should provide a reasonable alternative because neonates demonstrate only a limited adaptive immune response that would allow them to generate IgG antibodies independently [58], and the maternal antibodies received through breastfeeding are IgA.

## Data Availability

We welcome collaboration. Please contact the principal investigators, Dr. Christopher D. Heaney and Dr. Alain B. Labrique, for more information.

## COLLABORATION

The PAIR Study was designed and conducted through collaboration among investigators at the Johns Hopkins University, Columbia University, and the University of North Carolina at Chapel Hill in the United States; the University of Graz in Austria; and the JiVitA Maternal and Child Health and Nutrition Research Project in Bangladesh. We welcome further collaboration. Please contact the principal investigators, Dr. Christopher D. Heaney and Dr. Alain B. Labrique, for more information.

## FURTHER DETAILS

### Contributions

LNA and TJSS contributed equally to this paper. **LNA:** Writing – Original Draft, Data Curation, Methodology, Visualization; **TJSS:** Writing – Original Draft, Data Curation, Formal Analysis, Software, Visualization; **ANA:** Writing – Review & Editing, Conceptualization, Methodology, Supervision; **KK:** Data Curation, Resources; **NP:** Writing – Review & Editing, Conceptualization, Data Curation, Methodology, Project Administration; **PRR:** Methodology, Project Administration; **B Detrick:** Writing – Review & Editing, Conceptualization, Methodology; **RCF:** Writing – Review & Editing, Conceptualization, Methodology; **AvG:** Writing – Review & Editing, Investigation, Methodology; **WG:** Writing – Review & Editing, Investigation, Methodology; **RAK:** Writing – Review & Editing, Conceptualization, Methodology; **SLK:** Writing – Review & Editing, Conceptualization, Methodology; **ELO:** Methodology; **MWK:** Conceptualization, Methodology, Resources; **K Alland:** Writing – Review & Editing, Conceptualization, Methodology, Supervision; **K Ayesha:** Investigation, Supervision; **B Dyer:** Data Curation, Software; **MTI:** Investigation, Methodology; **HAO:** Investigation, Project Administration; **MHR:** Investigation, Methodology, Supervision; **HA:** Investigation, Methodology, Project Administration, Supervision; **RH:** Data Curation, Methodology, Supervision; **SS:** Conceptualization, Methodology, Project Administration, Supervision; **KJS:** Writing – Review & Editing, Conceptualization; **AKMM:** Investigation, Methodology; **ASMA:** Conceptualization, Methodology, Supervision; **MSF:** Methodology, Supervision; **KPW:** Conceptualization, Funding Acquisition, Supervision; **ABL:** Conceptualization, Funding Acquisition, Investigation, Methodology, Project Administration, Supervision; **CDH:** Writing – Review & Editing, Conceptualization, Data Curation, Funding Acquisition, Investigation, Methodology, Project Administration, Resources, Supervision.

## Ethics Approval

This study was approved by the institutional review boards of the Johns Hopkins Bloomberg School of Public Health (00008247) and the Institute of Epidemiology, Disease Control and Research (IEDCR/IRB/2017/07). All participants gave informed consent prior to enrollment.

## REFERENCES

1 Ravenscroth P, Brammer H, Richards K. Arsenic Pollution: A Global Synthesis. West Sussex: Wiley-Blackwell 2009. doi:10.1002/9781444308785

2 International Agency for Research on Cancer (IARC). Arsenic and Arsenic Compounds. Geneva: World Health Organization 2012.

3 Moon KA, Oberoi S, Barchowsky A, et al. A dose-response meta-analysis of chronic arsenic exposure and incident cardiovascular disease. Int J Epidemiol 2017;46:1924–39. doi:10.1093/ije/dyx202

4 Navas-Acien A, Spratlen MJ, Abuawad A, et al. Early-Life Arsenic Exposure, Nutritional Status, and Adult Diabetes Risk. Curr Diab Rep 2019;19:147–54. doi:10.1007/s11892-019-1272-9

5 Ahmed S, Moore SE, Kippler M, et al. Arsenic exposure and cell-mediated immunity in pre-school children in rural Bangladesh. Toxicol Sci 2014;141:166–75. doi:10.1093/toxsci/kfu113

6 Welch BM, Branscum A, Geldhof GJ, et al. Evaluating the effects between metal mixtures and serum vaccine antibody concentrations in children: A prospective birth cohort study. Environ Health 2020;19:41. doi:10.1186/s12940-020-00592-z

7 Welch BM, Branscum A, Ahmed SM, et al. Arsenic exposure and serum antibody concentrations to diphtheria and tetanus toxoid in children at age 5: A prospective birth cohort in Bangladesh. Environ Int 2019;127:810–8. doi:10.1016/j.envint.2019.04.015

8 Raqib R, Ahmed S, Ahsan K bin, et al. Humoral immunity in arsenic-exposed children in rural Bangladesh: Total immunoglobulins and vaccine-specific antibodies. Environ Health Perspect 2017;125:067006. doi:10.1289/EHP318

9 Heaney CD, Kmush B, Navas-Acien A, et al. Arsenic exposure and hepatitis E virus infection during pregnancy. Environ Res 2015;142:273–80. doi:10.1016/j.envres.2015.07.004

10 Kile ML, Rodrigues EG, Mazumdar M, et al. A prospective cohort study of the association between drinking water arsenic exposure and self-reported maternal health symptoms during pregnancy in Bangladesh. Environ Health 2014;13. doi:10.1186/1476-069X-13-29

11 Smith AH, Marshall G, Yuan Y, et al. Evidence from Chile that arsenic in drinking water may increase mortality from pulmonary tuberculosis. Am J Epidemiol 2011;173:414–20. doi:10.1093/aje/kwq383

12 Smith AH, Marshall G, Yuan Y, et al. Increased mortality from lung cancer and bronchiectasis in young adults after exposure to arsenic in utero and in early childhood. Environ Health Perspect 2006;114:1293–6. doi:10.1289/ehp.8832

13 Attreed SE, Navas-Acien A, Heaney CD. Arsenic and Immune Response to Infection During Pregnancy and Early Life. Cur Environ Health Rep 2017;4:229–43. doi:10.1007/s40572-017-0141-4

14 Islam LN, Nabi AHMN, Rahman MM, et al. Association of respiratory complications and elevated serum immunoglobulins with drinking water arsenic toxicity in human. J Environ Sci Health A Tox Hazard Subst Environ Eng 2007;42:1807–14. doi:10.1080/10934520701566777

15 Farzan SF, Korrick S, Li Z, et al. In utero arsenic exposure and infant infection in a United States cohort: a prospective study. Environ Res 2013;126:24–30. doi:10.1016/j.envres.2013.05.001

16 Farzan SF, Li Z, Korrick SA, et al. Infant Infections and Respiratory Symptoms in Relation to in Utero Arsenic Exposure in a U.S. Cohort. Environ Health Perspect 2016;124:840–7. doi:10.1289/ehp.1409282

17 Ahmed SM, Branscum A, Welch BM, et al. A prospective cohort study of in utero and early childhood arsenic exposure and infectious disease in 4-to 5-year-old Bangladeshi children. Environ Epidemiol 2020;4:e086. doi:10.1097/EE9.0000000000000086

18 George CM, Brooks WA, Graziano JH, et al. Arsenic exposure is associated with pediatric pneumonia in rural Bangladesh: a case control study. Environ Health 2015;14:83. doi:10.1186/s12940-015-0069-9

19 Rahman A, Vahter M, Ekström E-C, et al. Arsenic exposure in pregnancy increases the risk of lower respiratory tract infection and diarrhea during infancy in Bangladesh. Environ Health Perspect 2011;119:719–24. doi:10.1289/ehp.1002265

20 Raqib R, Ahmed S, Sultana R, et al. Effects of in utero arsenic exposure on child immunity and morbidity in rural Bangladesh. Toxicol Lett 2009;185:197–202. doi:10.1016/j.toxlet.2009.01.001

21 Smith AH, Yunus M, Khan AF, et al. Chronic respiratory symptoms in children following in utero and early life exposure to arsenic in drinking water in Bangladesh. Int J Epidemiol 2013;42:1077–86. doi:10.1093/ije/dyt120

22 Kuo CC, Moon KA, Wang SL, et al. The association of arsenic metabolism with cancer, cardiovascular disease, and diabetes: A systematic review of the epidemiological evidence. Environ Health Perspect 2017;125. doi:10.1289/EHP577

23 Hopenhayn C, Huang B, Christian J, et al. Profile of urinary arsenic metabolites during pregnancy. Environ Health Perspect 2003;111:1888–91. doi:10.1289/ehp.6254

24 Concha G, Vogler G, Lezcano D, et al. Exposure to inorganic arsenic metabolites during early human development. Toxicol Sci 1998;44:185–90. doi:10.1006/toxs.1998.2486

25 World Health Organization. WHO position paper on influenza vaccines - November 2012. Weekly Epidemiological Record 2012;87:461–76.

26 Zaman K, Roy E, Arifeen SE, et al. Effectiveness of maternal influenza immunization in mothers and infants. New Engl J Med 2008;359:1555–64. doi:10.1056/NEJMoa0708630

27 Reuman PD, Ayoub EM, Small PA. Effect of passive maternal antibody on influenza illness in children: a prospective study of influenza A in mother-infant pairs. Pediatr Infect Dis J 1987;6:398–403. doi:10.1097/00006454-198704000-00011

28 Puck JM, Glezen WP, Frank AL, et al. Protection of infants from infection with influenza A virus by transplacentally acquired antibody. J Infect Dis 1980;142:844–9. doi:10.1093/infdis/142.6.844

29 Glezen WP, Taber LH, Frank AL, et al. Influenza virus infections in infants. Pediatr Infect Dis J 1997;16:1065–8. doi:10.1097/00006454-199711000-00012

30 Neuzil KM, Reed GW, Mitchel EF, et al. Impact of influenza on acute cardiopulmonary hospitalizations in pregnant women. Am J Epidemiol 1998;148:1094–102. doi:10.1093/oxfordjournals.aje.a009587

31 Ser PH, Banu B, Jebunnesa F, et al. Arsenic exposure increases maternal but not cord serum IgG in Bangladesh. Pediatr Int 2015;57:119–25. doi:10.1111/ped.12396

32 Attreed SE, Navas-Acien A, Heaney CD. Arsenic and Immune Response to Infection During Pregnancy and Early Life. Curr Environ Health Rep 2017;4:229–43. doi:10.1007/s40572-017-0141-4

33 Flanagan S v, Johnston RB, Zheng Y. Arsenic in tube well water in Bangladesh: health and economic impacts and implications for arsenic mitigation. Bull World Health Organ 2012;90:839–46. doi:10.2471/BLT.11.101253

34 Ali H, Hamadani J, Mehra S, et al. Effect of maternal antenatal and newborn supplementation with Vitamin A on cognitive development of school-aged children in rural Bangladesh: A follow-up of a placebo-controlled, randomized trial. Am J Clin Nutr 2017;106:77–87. doi:10.3945/ajcn.116.134478

35 Klemm RDW, Labrique AB, Christian P, et al. Newborn Vitamin A Supplementation Reduced Infant Mortality in Rural Bangladesh. Pediatrics 2008;122:e242–50. doi:10.1542/peds.2007-3448

36 West Jr. KP, Christian P, Labrique AB, et al. Effects of vitamin A or beta carotene supplementation on pregnancy-related mortality and infant mortality in rural Bangladesh: A cluster randomized trial. JAMA 2011;305:1986–95. doi:10.1001/jama.2011.656

37 West Jr. KP, Shamim AA, Mehra S, et al. Effect of Maternal Multiple Micronutrient vs Iron-Folic Acid Supplementation on Infant Mortality and Adverse Birth Outcomes in Rural Bangladesh: The JiVitA-3 Randomized Trial. JAMA 2014;312:2649–58. doi:10.1001/jama.2014.16819

38 Serradell L, Wagué S, Moureau A, et al. Enhanced passive safety surveillance of a trivalent and a quadrivalent influenza vaccine in Denmark and Finland during the 2018/2019 season. Hum Vaccin Immunother 2021;17:1205–10. doi:10.1080/21645515.2020.1804247

39 Gunnsteinsson S, Labrique AB, West KP, et al. Constructing indices of rural living standards in Northwestern Bangladesh. J Health Popul Nutr 2010;28:509–19. doi:10.3329/jhpn.v28i5.6160

40 Cheng Z, Zheng Y, Mortlock R, et al. Rapid multi-element analysis of groundwater by high-resolution inductively coupled plasma mass spectrometry. Anal Bioanal Chem 2004;379:512–8. doi:10.1007/s00216-004-2618-x

41 Weinberg A. Cryopreservation of Peripheral Blood Mononuclear Cells. In: Detrick B, Schmitz JL, Hamilton RG, eds. Manual of Molecular and Clinical Laboratory Immunology. Washington, DC: ASM Press 2016. 263–8.

42 Ohmit SE, Petrie JG, Cross RT, et al. Influenza hemagglutination-inhibition antibody titer as a correlate of vaccine-induced protection. J Infect Dis 2011;204:1879–85. doi:10.1093/infdis/jir661

43 Scheer J, Findenig S, Goessler W, et al. Arsenic species and selected metals in human urine: validation of HPLC/ICPMS and ICPMS procedures for a long-term population-based epidemiological study. Anal Methods 2012;4:406–13. doi:10.1039/C2AY05638K

44 National Research Council. Arsenic in Drinking Water. Washington, DC:: National Academies Press 1999. doi:10.17226/6444

45 McKie A, Vyse A, Maple C. Novel methods for the detection of microbial antibodies in oral fluid. Lancet Infect Dis 2002;2:18–24. doi:10.1016/s1473-3099(01)00169-4

46 Randad PR, Hayford K, Baldwin R, et al. The Utility of Antibodies in Saliva to Measure Pathogen Exposure and Infection. In: Granger Douglas A., Taylor Marcus K., eds. Salivary Bioscience: Foundations of Interdisciplinary Saliva Research and Applications. Cham, Switzerland: Springer Nature 2020. 287–319. doi:10.1007/978-3-030-35784-9

47 Brandtzaeg PER. Do salivary antibodies reliably reflect both mucosal and systemic immunity? Ann N Y Acad Sci 2007;1098:288–311. doi:10.1196/annals.1384.012

48 Randad PR, Pisanic N, Kruczynski K, et al. COVID-19 serology at population scale: SARS-CoV-2-specific antibody responses in saliva. J Clin Microbiol 2020;59:e02204. doi:10.1128/JCM.02204-20

49 Fitzner J, Qasmieh S, Mounts AW, et al. Revision of clinical case definitions: influenza-like illness and severe acute respiratory infection. Bull World Health Organ 2018;96:122–8. doi:10.2471/BLT.17.194514

50 World Health Organization (WHO). Information for the Molecular Detection of Influenza Viruses. 2017.https://www.who.int/influenza/gisrs_laboratory/WHO_information_for_the_molecular_detection_of_influenza_viruses_20171023_Final.pdf

51 West Jr. KP, Shamim AA, Labrique AB, et al. Efficacy of Antenatal Multiple Micronutrient (MM) vs Iron-Folic Acid (IFA) Supplementation in Improving Gestational and Postnatal Viability in Rural Bangladesh: The JiVitA-3 Trial. FASEB J 2013;27:358.6-358.6. doi:10.1096/fasebj.27.1_supplement.358.6

52 Kim JM, Labrique A, West KP, et al. Maternal morbidity in early pregnancy in rural northern Bangladesh. Int J Gynaecol Obstet 2012;119:227–33. doi:10.1016/j.ijgo.2012.06.022

53 World Health Organization (WHO). Haemoglobin concentrations for the diagnosis of anaemia and assessment of severity. Geneva, Switzerland: 2011. https://www.who.int/vmnis/indicators/haemoglobin.pdf

54 Ahsan H, Chen Y, Parvez F, et al. Health Effects of Arsenic Longitudinal Study (HEALS): description of a multidisciplinary epidemiologic investigation. J Expo Sci Environ Epidemiol 2006;16:191–205. doi:10.1038/sj.jea.7500449

55 Rahman A, Vahter M, Ekström E-C, et al. Association of arsenic exposure during pregnancy with fetal loss and infant death: a cohort study in Bangladesh. Am J Epidemiol 2007;165:1389–96. doi:10.1093/aje/kwm025

56 Navas-Acien A, Francesconi KA, Silbergeld EK, et al. Seafood intake and urine concentrations of total arsenic, dimethylarsinate and arsenobetaine in the US population. Environmental Research 2011;111:110–8. doi:10.1016/j.envres.2010.10.009

57 Huhmann LB, Harvey CF, Navas-Acien A, et al. A mass-balance model to assess arsenic exposure from multiple wells in Bangladesh. J Expo Sci Environ Epidemiol 2021;:1–9. doi:10.1038/s41370-021-00387-5

58 Semmes EC, Chen J-L, Goswami R, et al. Understanding early-life adaptive immunity to guide interventions for pediatric health. Frontiers in Immunology 2021;11:3544. doi:10.3389/fimmu.2020.595297

